# Automated assessment of COVID-19 pulmonary disease severity on chest radiographs using convolutional Siamese neural networks

**DOI:** 10.1101/2020.05.20.20108159

**Authors:** Matthew D. Li, Nishanth Thumbavanam Arun, Mishka Gidwani, Ken Chang, Francis Deng, Brent P. Little, Dexter P. Mendoza, Min Lang, Susanna I. Lee, Aileen O’Shea, Anushri Parakh, Praveer Singh, Jayashree Kalpathy-Cramer

## Abstract

**Purpose:** To develop an automated measure of COVID-19 pulmonary disease severity on chest radiographs (CXRs), for longitudinal disease evaluation and clinical risk stratification.

**Materials and Methods:** A convolutional Siamese neural network-based algorithm was trained to output a measure of pulmonary disease severity on anterior-posterior CXRs (pulmonary x-ray severity (PXS) score), using weakly-supervised pretraining on ~160,000 images from CheXpert and transfer learning on 314 CXRs from patients with COVID-19. The algorithm was evaluated on internal and external test sets from different hospitals, containing 154 and 113 CXRs respectively. The PXS score was correlated with a radiographic severity score independently assigned by two thoracic radiologists and one in-training radiologist. For 92 internal test set patients with follow-up CXRs, the change in PXS score was compared to radiologist assessments of change. The association between PXS score and subsequent intubation or death was assessed.

**Results:** The PXS score correlated with the radiographic pulmonary disease severity score assigned to CXRs in the COVID-19 internal and external test sets (ρ=0.84 and ρ=0.78 respectively). The direction of change in PXS score in follow-up CXRs agreed with radiologist assessment (ρ=0.74). In patients not intubated on the admission CXR, the PXS score predicted subsequent intubation or death within three days of hospital admission (area under the receiver operator characteristic curve=0.80 (95%CI 0.75-0.85)).

**Conclusion:** A Siamese neural network-based severity score automatically measures COVID-19 pulmonary disease severity in chest radiographs, which can be scaled and rapidly deployed for clinical triage and workflow optimization.

**SUMMARY:** A convolutional Siamese neural network-based algorithm can calculate a continuous radiographic pulmonary disease severity score in COVID-19 patients, which can be used for longitudinal disease evaluation and clinical risk stratification.

**KEY RESULTS:**

- A Siamese neural network-based severity score correlates with radiologist-annotated pulmonary disease severity on chest radiographs from patients with COVID-19 (ρ=0.84 and ρ=0.78 in internal and external test sets respectively).
- The direction of change in the severity score in follow-up radiographs is concordant with radiologist assessment (ρ=0.74).
- The admission chest radiograph severity score can help predict subsequent intubation or death within three days of admission (receiver operator characteristic area under the curve=0.80).

## INTRODUCTION

The role of diagnostic chest imaging continues to evolve during the COVID-19 pandemic. According to American College of Radiology guidelines, while chest CT is not recommended for COVID-19 diagnosis or screening, portable chest radiographs (CXRs) are suggested when medically necessary (1). The Fleischner Society has stated that CXRs can be useful for assessing COVID-19 disease progression (2) and one study found that 69% of these patients have an abnormal baseline CXR (3).

While radiographic findings are neither sensitive nor specific for COVID-19, with findings overlapping other infections and pulmonary edema, CXRs can be useful for assessing pulmonary infection severity and evaluating longitudinal changes. However, there is substantial variability in the interpretations of CXRs by radiologists, as has been demonstrated for pneumonia (4–6). In addition, commonly used disease severity categories on chest radiographs, such as “mild,” “moderate,” and “severe,” are challenging to reproduce as the thresholds between these categories are subjective.

One possible solution to these challenges is to train a convolutional Siamese neural network to estimate radiographic disease severity on a continuous spectrum (schematic in Figure 1A) (7). Siamese neural networks take two separate images as inputs, which are passed through twinned neural networks (8,9). The Euclidean distance between the final two layers of the networks can be calculated, which serves as a measure of distance between the two images with respect to the imaging features being trained on, such as disease features. If an image-of-interest is compared pairwise to a pool of “normal” images, the disease severity can be abstracted to the median of those Euclidean distances.

**Figure 1:**
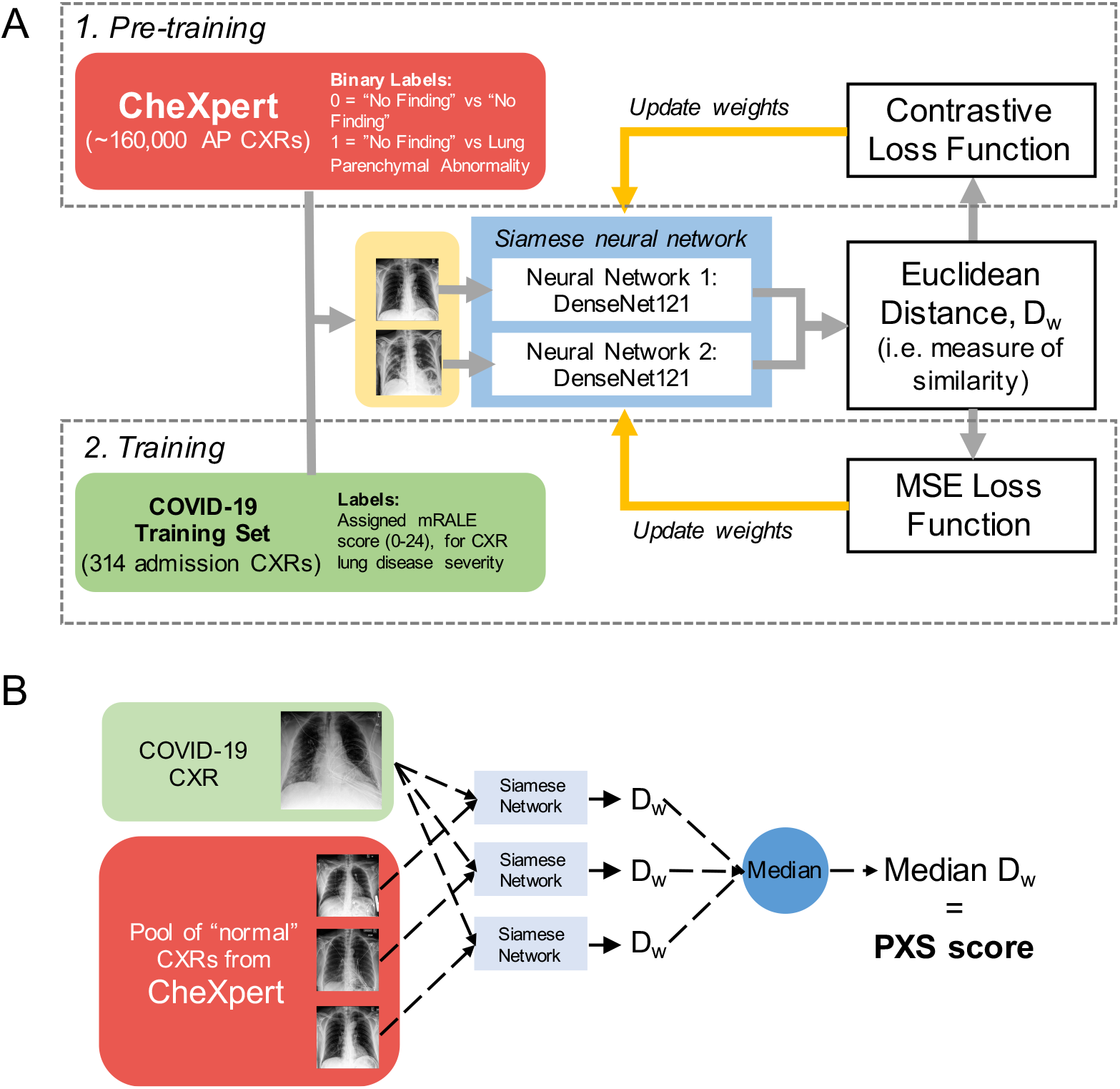
A, Schematic for training the convolutional Siamese neural network-based algorithm used to calculate the Pulmonary X-Ray Severity (PXS) score, a continuous measure of radiographic pulmonary disease severity in COVID-19 patients. The network is pre-trained with chest radiographs (CXRs) from CheXpert (9) using binary lung disease presence labels and then trained on CXRs from a COVID-19 training set using annotations for modified Radiographic Assessment of Lung Edema (mRALE) scores. B, Schematic for calculating the PXS score, which is calculated by comparing the image of interest pairwise with a pool of normal CXRs from CheXpert. D_w_ = Euclidean distance; MSE loss = mean square error loss.

In this study, we hypothesized that a convolutional Siamese neural network-based algorithm could be trained to yield a measure of radiographic pulmonary disease severity on anterior-posterior (AP) CXRs (pulmonary x-ray severity (PXS) score). We evaluated the algorithm performance on internal and external test sets of CXRs from patients with COVID-19. We also investigated the association between the admission PXS score and subsequent intubation or death.

## MATERIALS AND METHODS

This Health Insurance Portability and Accountability Act-compliant retrospective study was reviewed and exempted by the Institutional Review Board of Massachusetts General Hospital, with waiver of informed consent.

### Public Chest Radiograph Data

We obtained a publicly available CXR data set, CheXpert, from Stanford Hospital, Palo Alto (10), containing 224,316 CXRs. Each image has labels for 14 radiographic observations extracted from the impression section of corresponding free-text radiology reports, derived from a rule-based algorithm that classifies entities as negative, uncertain, or positive. For the purpose of training our deep learning algorithm, we grouped together uncertain and positive labels together as positive labels, as uncertainty still implies an image abnormality.

This data set also includes annotations for image view, which we used to filter for AP radiographs only, as suspected or confirmed COVID-19 positive patients tend to be imaged more frequently in the AP projection in emergency room and hospital settings. CheXpert includes a partition for training and validation, and after filtering for only AP images, the training and validation sets contained 161,590 and 169 images, respectively.

### COVID-19 Positive Chest Radiograph Data

We obtained raw DICOM data for CXRs at a large urban quaternary-care hospital in the United States (Massachusetts General Hospital, Boston, MA), from COVID-19 positive patients (confirmed by nasopharyngeal swab RT-PCR). The COVID-19 training set contained 314 admission CXRs from unique patients hospitalized at least in part from April 1-10, 2020, randomly partitioned 9:1 for training and validation (282:32 images). The COVID-19 internal test set contained 154 admission CXRs from unique patients hospitalized at least in part from March 27-31, 2020. One patient with COVID-19 was omitted due to a pneumonectomy. There was no overlap between training and test set patients. Among the COVID-19 internal test set patients, 92 underwent a follow-up CXR within 12 days of admission. The DICOM data for these follow-up radiographs were also obtained. For DICOMs containing more than one frontal image acquisition, the standard CXR image without postprocessing was selected, with the best positioning available. Intubation and mortality data were collected from the medical record by two investigators blinded to CXR findings. We also obtained raw DICOM data for 113 admission CXRs associated with unique patients hospitalized at least in part on April 15, 2020 at a community hospital in the United States (Newton Wellesley Hospital, Newton, MA), from COVID-19 positive patients (confirmed by nasopharyngeal swab RT-PCR), which served as an external test set.

### Radiologist Scoring of Pulmonary Disease Severity on Chest Radiographs

To provide a reference standard assessment of disease severity on CXRs, we used a simplified version of the Radiographic Assessment of Lung Edema (RALE) score (11). This grading scale was originally validated for use in pulmonary edema assessment in acute respiratory distress syndrome (ARDS) and incorporates the extent and density of alveolar opacities on CXRs. The grading system is relevant to COVID-19 patients as the CXR findings tend to involve multifocal alveolar opacities (3) and many hospitalized COVID-19 patients develop ARDS (12). In our study, we use a modified RALE (mRALE) score. Each lung is assigned a score for the extent of involvement by consolidation or ground glass/hazy opacities (0=none; 1=<25%; 2=25-50%; 3=50-75%; 4=>75% involvement). Each lung score is then multiplied by an overall density score (1=hazy, 2=moderate, 3=dense). The sum of scores from each lung is the mRALE score. Thus, a normal CXR receives a score of 0, while a CXR with complete consolidation of both lungs receives the maximum score of 24. mRALE differs from the original RALE score in that the lungs are not divided into quadrants.

Using the mRALE scoring system, two in-training radiologists (M.D.L. and F.D., both postgraduate year 4) annotated each image in the COVID-19 training set, independently viewing the post-processed JPEG images. Two fellowship-trained thoracic radiologists (B.P.L., 11 years of experience; D.P.M., 2 years of experience) and an in-training radiologist (M.D.L. for the internal test set and F.D. for the external test set) annotated each image in the COVID-19 internal and external test sets, independently viewing the CXRs in a PACS system. The reference standard mRALE score for each image is the average of the raters.

### Radiologist Assessment of Longitudinal Change

The same raters who assessed the COVID-19 internal test set also evaluated the 92 internal test set patients with follow-up CXRs. For each longitudinal image pair (displayed as side-by-side post-processed JPEG images), the raters independently assigned the label: decreased, same, or increased pulmonary disease severity. The majority change label was assigned with two or more votes for one label.

### Convolutional Siamese Neural Network Training

A convolutional Siamese neural network architecture takes two separate images as inputs, which are separately passed through identical subnetworks with shared weights (schematic in Figure 1A, see Supplemental Materials for image pre-processing details) (8,9). We built such a network using DenseNet121 (13) as the underlying subnetwork with initial pre-training on ImageNet, as this architecture had empirically performed well for classification tasks in the CheXpert study (10). The Euclidean distance D_w_ between the subnetwork outputs, G_w_(X_1_) and G_w_(X_2_), given image input vectors X_1_ and X_2_, is calculated from the equation (*D_w_(X_1_,X*_2_) = ||*G_w_(Z*_1_) − *G_w_*(X_2_)||_2_) (9).

We used a two-step training strategy, that involves pre-training with weak labels on the large CheXpert data set using the contrastive loss function (8), followed by transfer learning to the relatively small COVID-19 training set using mean square error loss, using the assigned mRALE scores as disease severity labels. Details regarding the training strategy are in the Supplemental Materials. The code is available at *https://github.com/QTIM-Lab/PXS-score*.

### Calculating the Pulmonary X-Ray Severity (PXS) Score

After training the Siamese neural network, when two CXR images are passed through the subnetworks, the Euclidean distance calculated from the subnetwork outputs can serve as a continuous measure of difference between the two CXRs, with respect to pulmonary parenchymal findings. Thus, to evaluate a single image-of-interest for pulmonary disease severity, an image can be compared to a pool of *N* images without a lung abnormality (schematic in Figure 1B). We empirically set *N*=12, using all cases labeled with “No Finding” from the CheXpert validation set as the normal pool (ages 19-68 years, 7 women). Using the Siamese neural network, the Euclidean distance is calculated between the image-of-interest and each of the *N* normal images, and the median Euclidean distance is calculated. This median Euclidean distance is the Pulmonary X-Ray Severity (PXS) score.

### Occlusion sensitivity maps for visualizing Siamese neural network outputs

We used an occlusion sensitivity approach (14) to visualize what portions of the input images were important to the Siamese neural network for calculating the PXS score. See the Supplemental Materials for details.

### Statistical Analysis

We used Chi-square and Mann-Whitney tests, Spearman rank correlation (ρ), linear Cohen’s kappa (κ), and bootstrap 95% confidence intervals where appropriate (see Supplementary Materials for details). The threshold for statistical significance was considered *a priori* to be P<0.05.

## RESULTS

### COVID-19 Patient Data Set Characteristics

In the COVID-19 training, internal test, and external test sets, the rank correlation between the mRALE scores assigned by the radiologist raters was similar (ρ=0.82-0.85, P<0.001 in all cases; see Supplemental Materials for details). Patients in the external test set were significantly older than patients in the training and internal test sets (median age 57 years versus 74 years respectively, P<0.001). There was no significant difference in gender between the two sites (48% versus 41% female respectively, P=0.2).

There were 468 unique patients in total from the COVID-19 training and internal test sets with available data on intubation or death. Of these patients, 134 were intubated or dead within 3 days of hospital admission (including 31 with an endotracheal tube on the admission CXR) and 334 were not intubated or dead. There was no significant difference in gender between patients who were intubated or dead and those who were not (43% versus 37% female respectively, P=0.4). Patients who were intubated or dead were slightly older than those who were not (median age 60 years (interquartile range 50-72) versus 56 years (interquartile range 42-72) respectively, P=0.049).

### Siamese Neural Network-based PXS Score Correlates with Radiographic Pulmonary Disease Severity

In the 154-patient COVID-19 internal test set, the Siamese neural network-based Pulmonary X-Ray Severity (PXS) score correlated with the average mRALE score assigned, which is a measure of radiographic pulmonary disease severity (ρ=0.84, P<0.001) (Figure 2A). In the 113-patient COVID-19 external test set, the PXS score also correlated with the average mRALE score assigned (ρ=0.78, P<0.001) (Figure 2B). Using an occlusion sensitivity map-based approach, we show that the network focuses its attention on pulmonary opacities (Figure 2C).

**Figure 2:**
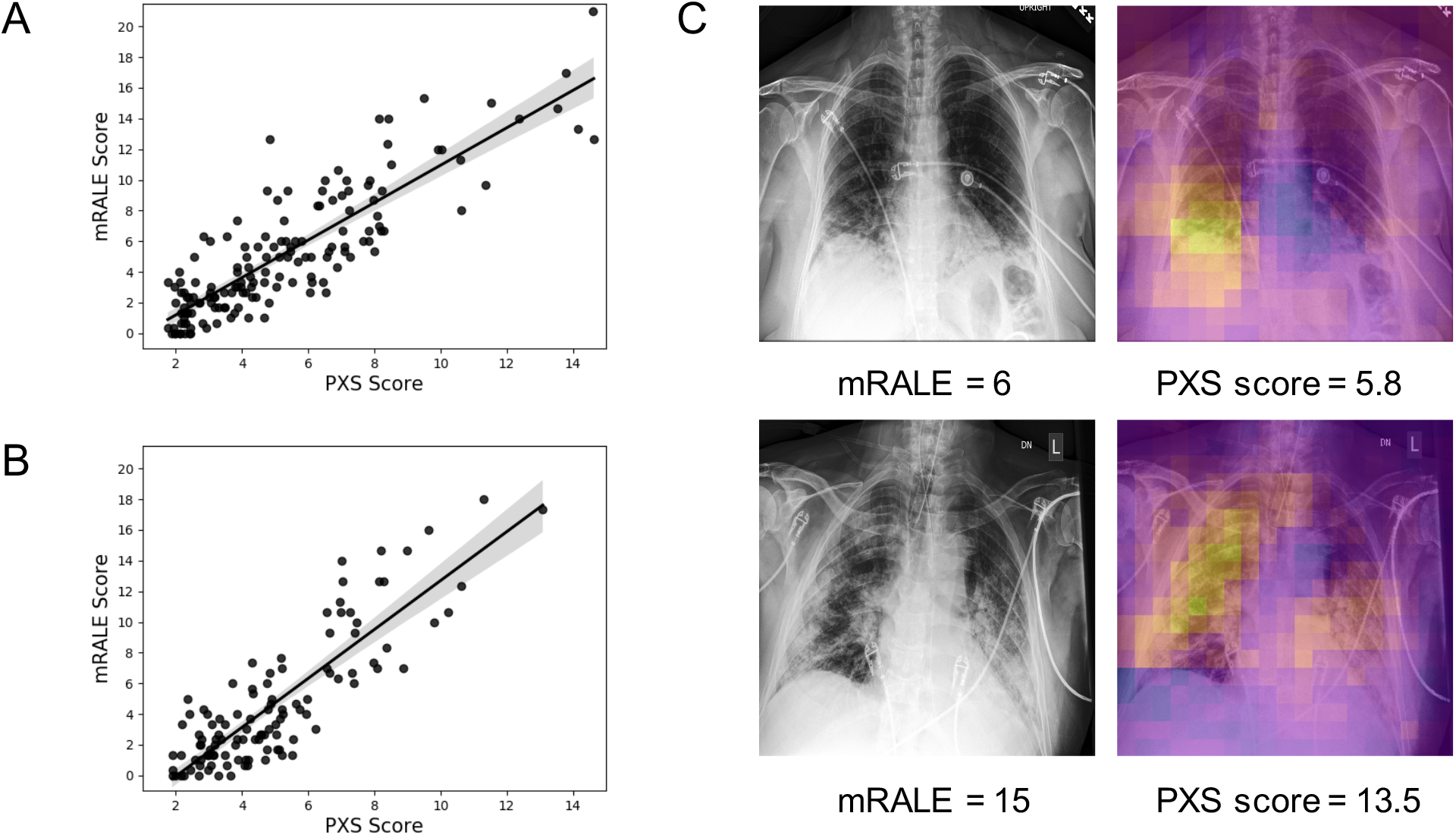
Siamese neural network-based Pulmonary X-Ray Severity (PXS) score is a measure of radiographic pulmonary disease severity in patients with COVID-19. A and B, Scatterplots show, in a 154-patient test set (A) and 113-patient external hospital test set (B), the PXS score correlates with the modified Radiographic Assessment of Lung Edema (mRALE) score, a measure of pulmonary disease severity on chest radiographs (ρ=0.84, P<0.001 and ρ=0.78, P<0.001, respectively) (linear regression 95% confidence interval shown in the scatterplots). C, Occlusion sensitivity map-based approach shows that the Siamese neural network is focusing on pulmonary opacities. Yellow areas indicate parts of the image important to the neural network.

We also trained a Siamese neural network using only the 314-image COVID-19 training set, without the weakly-supervised pre-training using the large ChexPert data set. The PXS score from this model had lower correlation with the mRALE score in the internal test set (ρ=0.81, P<0.001) and external test set (ρ=0.77, P<0.001), demonstrating a slight performance boost from pre-training.

### Longitudinal Change Assessment with the PXS Score

Of the COVID-19 internal test set patients with available longitudinal CXRs, according to the assigned majority vote change labels, 24 (26%), 19 (21%), and 44 (48%) of patients showed a decrease, no change, or increase in pulmonary disease severity respectively. Five patients (5%) did not receive majority votes (i.e. the three raters each voted differently) and were omitted from further analysis, which reflects subjectivity in the interpretation of heterogeneous CXRs. The inter-rater reliability between the three raters for assigning change labels was moderate (linear Cohen’s κ = 0.58, 0.59, 0.57).

The change in PXS score between two longitudinally acquired images correlates with the majority vote change label (ρ=0.74, P<0.001) (Figure 3A). For patients labeled with decreased disease severity, 18 (75%) were associated with decreased PXS score. For patients labeled for increased disease severity, 43 (98%) were associated with increased PXS score. For patients labeled for no change, the mean PXS score change is 0.1 (standard deviation ± 1.3). Illustrative examples of longitudinal change assessment are shown in Figure 3B.

**Figure 3:**
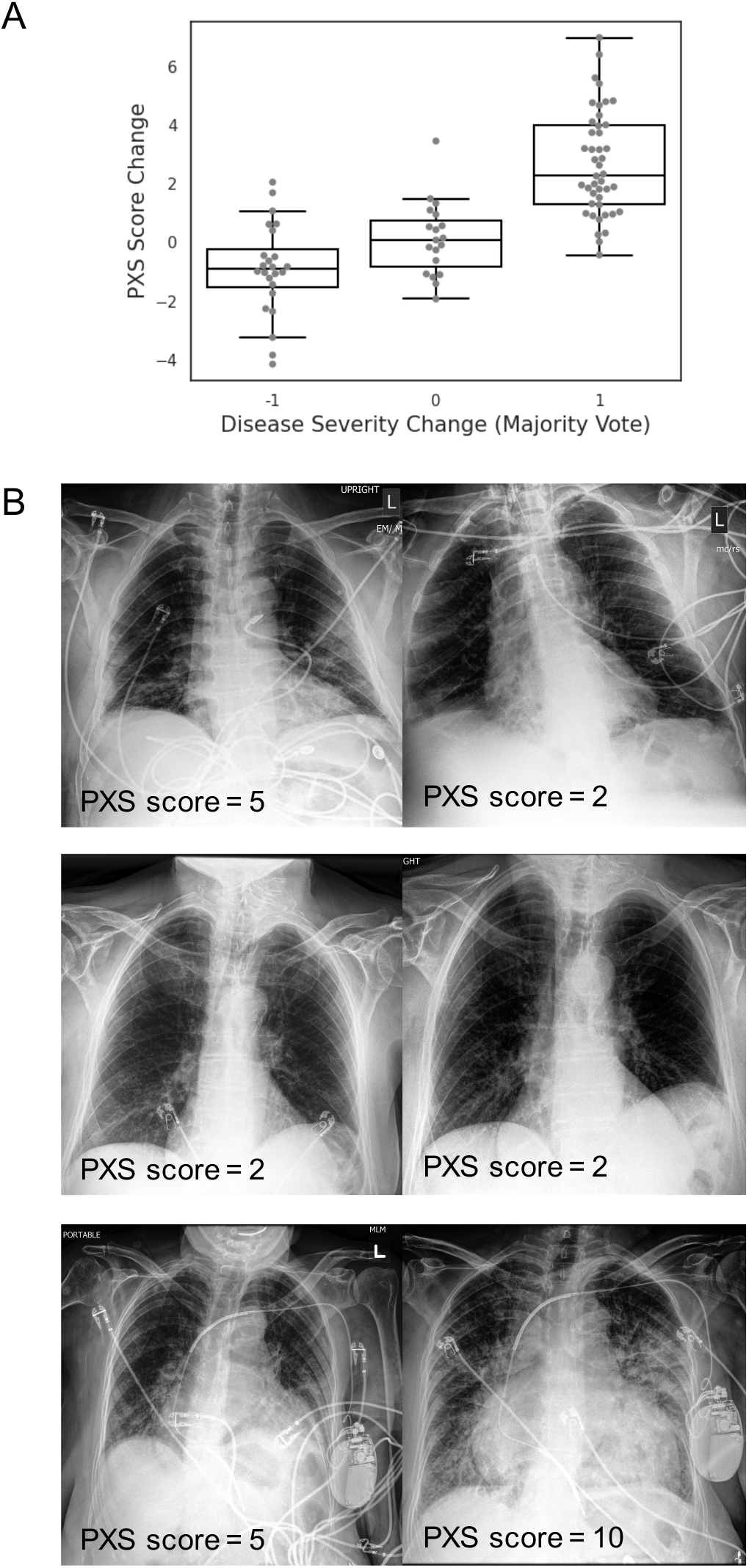
Siamese neural network-based Pulmonary X-Ray Severity (PXS) score can be used to assess longitudinal change in radiographic disease severity over time in COVID-19 patients. A, Boxplot shows the PXS score correlates with majority vote change in pulmonary disease severity (ρ=0.74, P<0.001), where -1, 0, and 1 indicate decreased, unchanged, and increased severity in longitudinal chest radiograph pairs, assigned by three independent raters (2 thoracic radiologists, 1 radiology resident). The boxplot boxes indicate the median and interquartile range (IQR), with whiskers extending to points within 1.5 IQRs of the IQR boundaries. B, Examples of PXS score evaluation of longitudinal change in three patients with COVID-19.

### Association Between PXS Score and Intubation or Death

The PXS score was significantly higher on admission CXRs of patients with COVID-19 who were intubated or dead within 3 days of admission from our training and internal test sets, compared to those who were not intubated (median PXS score 7.9 versus 3.2, P<0.001) (Figure 4A). Importantly, the PXS score algorithm is not trained on outcomes data. Of the 134 patients who were intubated or died within 3 days of admission, 76 were intubated on the admission day and 31, 12 and 14 patients on hospital days 1, 2, and 3 respectively. A higher PXS score is associated with a shorter time interval before intubation or death in these patients (ρ=0.25, P=0.004) (Figure 4B).

**Figure 4:**
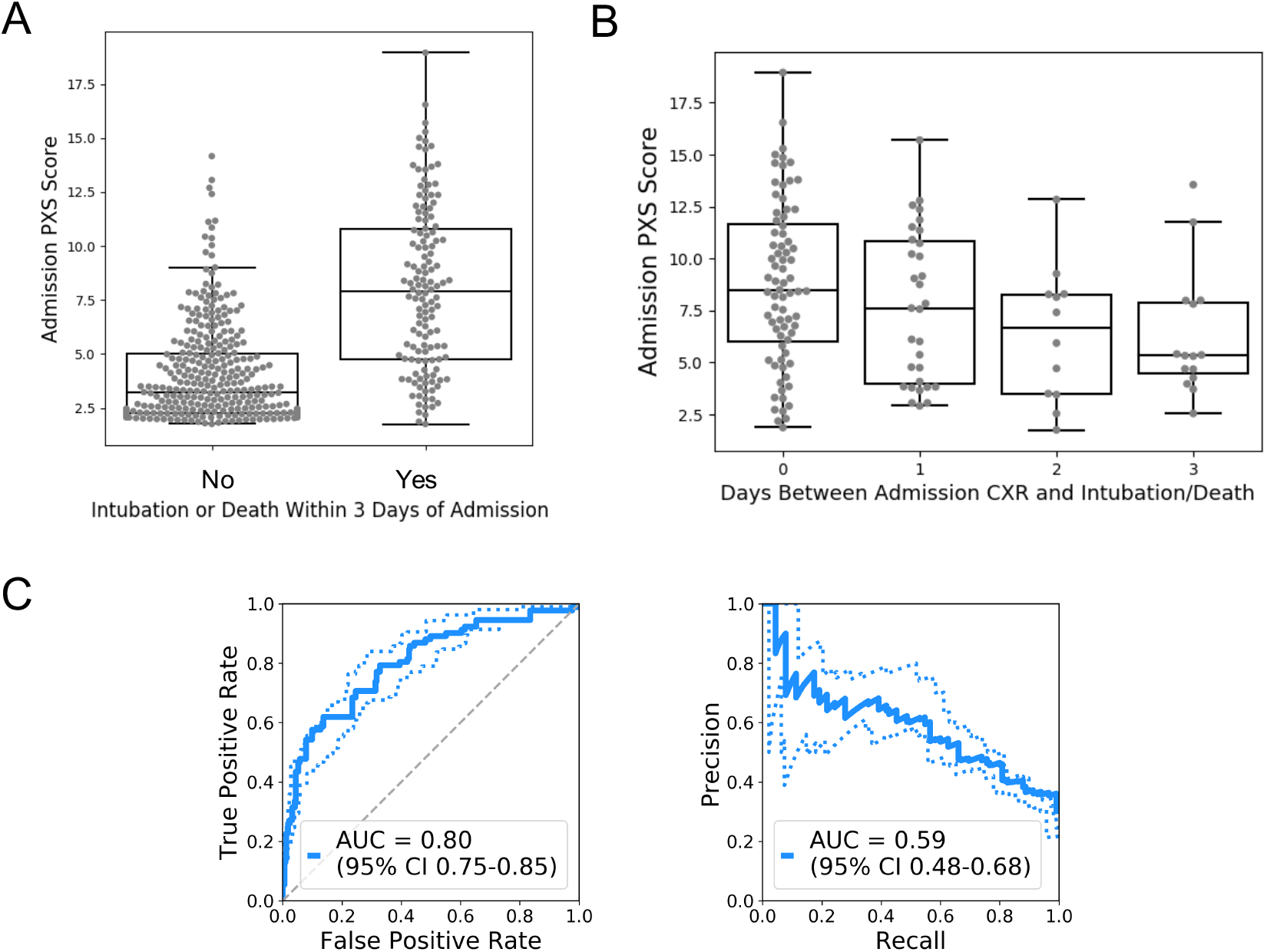
Siamese neural network-based Pulmonary X-Ray Severity (PXS) score is associated with intubation/death in patients hospitalized with COVID-19. A, Boxplot shows the PXS score is significantly higher in patients intubated or dead within three days of hospital admission (P<0.001). B, Boxplot shows that a higher PXS score is associated with a shorter time interval before intubation or death (ρ=0.25, P=0.004), CB, Receiver operator characteristic and precision recall curves show the performance of the PXS score for predicting subsequent intubation or death within three days of hospital admission, in patients without an endotracheal tube on their admission chest radiograph (AUC, area under the curve; dashed lines indicate bootstrap 95% confidence intervals).

Given these findings, we used the PXS score as a continuous input for prediction of intubation or death within 3 days of hospital admission. For the 437 patients without an endotracheal tube present on the admission CXR, the receiver operator characteristic area under the curve (AUC) was 0.80 (bootstrap 95% confidence interval 0.75-0.85) (Figure 4C). If the PXS score threshold is set to 3.1 for predicting intubation or death within 3 days, the sensitivity is 90% and specificity is 47% for these outcomes.

## DISCUSSION

Front-line clinicians estimate the risk for clinical decompensation in patients with COVID-19 using a combination of data, including epidemiologic factors, comorbidities, vital signs, lab values, and clinical intuition (12,15). The chest radiograph can help contribute to this assessment, but manual assessment of severity is subjective and requires expertise. In this study, we designed and trained a Siamese neural network-based algorithm to provide an automated measure of COVID-19 disease severity on chest radiographs in hospitalized patients, the Pulmonary X-ray Severity (PXS) score. The PXS score correlates with a manually annotated measure of radiographic disease severity in internal and external test sets, and the direction of change in PXS score for longitudinally acquired radiographs is concordant with radiologist assessment. For patients with COVID-19 presenting to the hospital with an admission chest radiograph, the PXS score can help predict subsequent intubation or death within three days of admission.

The automatic PXS score can potentially be rapidly scaled and deployed, which has important clinical applications in the COVID-19 pandemic, particularly in countries like the United States or under-resourced settings where CXRs are frequently acquired, while CT studies are relatively rarely obtained. The PXS score can assist with clinical decision making. For example, in the emergency room or urgent care setting, clinicians must decide whether or not a patient is safe to discharge home or requires hospital admission. By setting the PXS score threshold in favor of sensitivity (90% sensitivity, 47% specificity) for prediction of intubation or death within 3 days, the score can be used to help with such decisions. Additionally, the PXS score can be incorporated into multivariable machine learning models that account for other clinical variables like vital signs, lab values, and co-morbidities. Other potential applications include radiologist workflow optimization, where CXRs with more severe findings can be interpreted earlier, and hospital resource management, where the PXS score can help with resource allocation (e.g. prediction of future ventilator need).

Various grading systems have been developed to measure respiratory disease severity on chest imaging, including for pulmonary edema in ARDS (11), severe acute respiratory infection (16), parainfluenza virus-associated infections (17), and pediatric pneumonia (18). These studies use manually annotated features from chest imaging to predict outcomes, such as mortality, need for intensive care, and other adverse events. However, barriers to adoption of these systems include inter-rater reliability and learning curve for users. Our automated Siamese neural network-based approach addresses these challenges.

Deep learning-based algorithms have been applied to CXRs extensively, but primarily for disease detection, such as for pneumonia and tuberculosis (19,20), as well as for COVID-19 localization on CXR images (21). However, due to the nature of chest radiography, there are limits to the sensitivity and specificity of this modality for COVID-19 detection (3). There is a relative paucity of research using deep learning for disease severity assessment on CXRs. Automated evaluation of pulmonary edema severity on CXRs has been explored using a deep learning model that incorporates ordinal regression of edema severity labels in training (no, mild, moderate, or severe edema) (22). These severity labels were extracted from associated radiology reports, but are inherently noisy given the variability in interpretation of the CXRs (23,24). This problem of noisy labels extends beyond pulmonary edema to any disease process where there is subjectivity in interpretation. Our Siamese neural network-based approach mitigates the label noise via transfer learning on data labeled with mRALE, a more fine-grained scoring system which showed high agreement between raters in our study.

There are several limitations to this study. First, the COVID-19 patients in this study were from urban areas of the United States, which may limit the external generalizability of this algorithm to other locations. However, given that the model was able to generalize to a second hospital (community hospital vs quaternary care center) with only a small decrease in performance, we believe the model should be robust and share the code for further research. Second, abnormal patient positioning and respiratory phase may affect the score. Variability resulting from these differences may impact the algorithm performance in evaluating subtle changes between CXRs. However, since the algorithm explicitly learns to assess radiographic disease severity, quality control by a radiologist is relatively simple as the radiologist can compare the PXS score to what is expected on sample studies. Third, our algorithm was trained for use on AP chest radiographs, as AP positioning is more common than posterior-anterior images among patients with COVID-19. This limits the generalizability of the algorithm model for posterior-anterior radiographs.

We developed an automated Siamese neural network-based pulmonary disease severity score for patients with COVID-19, with the potential to help with clinical triage and workflow optimization. With further validation, the score could be incorporated into clinical treatment guidelines to be used together with other clinical and lab data. Beyond the COVID-19 pandemic, this automated severity score could also be modified and applied to other continuous disease processes manifesting on chest radiographs, like pulmonary edema, interstitial lung disease, and other infections.

## Data Availability

The CheXpert data is available through the Stanford ML Group (https://stanfordmlgroup.github.io/competitions/chexpert/). Patient data used in this study is not publicly available.

## ACKNOWLEDGMENTS

We thank Jeremy Irvin for sharing his DICOM pre-processing code for the CheXpert data set. We appreciate the work of our clinical colleagues and other frontline staff during the COVID-19 pandemic.

## Abbreviations

COVID-19: coronavirus disease 2019
CXR: chest radiograph
RT-PCR: reverse transcriptase-polymerase chain reaction
AP: anterior-posterior
mRALE score: modified Radiographic Assessment of Lung Edema score
PXS score: pulmonary x-ray severity score
AUC: area under the curve.

## SUPPLEMENTAL MATERIALS

### Supplemental Methods

#### Chest Radiograph Image Pre-processing

Full size CXR images in JPEG format from CheXpert were all resized to 320 x 320 pixels, which is within the resolution range of optimal performance for CXR binary classification tasks (25). DICOM files from the COVID-19 CXRs were all pre-processed in the same manner as in ChexPert, with image pixel array extraction using pydicom (26), followed by normalization to [0, 255], conversion to 8-bit, correction of photometric inversion, histogram equalization in OpenCV (27), and conversion to a JPEG file. In the external test set CXR images, some images included a large black border around the actual radiograph, which was mostly removed using an automatic cropping algorithm in Python (border pixels with a 0 pixel value were removed).

#### Convolutional Siamese Neural Network Training

We used a two-step training strategy, that involves pre-training with weak labels on the large CheXpert data set followed by transfer learning to the relatively small COVID-19 training set, as follows:

**Step 1**. To pre-train the Siamese neural network on CheXpert data, the contrastive loss function is used to train the network parameters, as defined by the equation 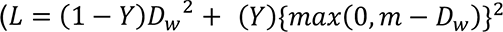; Y = 0 if same class (i.e. no change) and Y = 1 if different class (i.e. change), D_w_ = Euclidean distance, and m = margin) (9). The contrastive loss function minimizes when there is a small Euclidean distance for no change and large Euclidean distance for change in class. The margin hyperparameter is empirically set to 50, which gives the maximum D_w_ for which dissimilar image input pairs will not contribute further to the loss, helping to stabilized training. As the goal of this algorithm is to generate a measure of disease severity, we trained the convolutional Siamese neural network to maximize Euclidean distance when the input images showed a difference in labels that identify lung parenchymal abnormalities. In the CheXpert data set, there are annotations that represent pulmonary parenchymal findings, including “lung opacity,” “lung lesion,” “consolidation,” “pneumonia,” “atelectasis,” and “edema.” If an image had any one of these labels, it was assigned an abnormal lung label. If an image did not have any one of those labels, it was assigned a normal lung label. In training the network, paired CXR images were sampled from the training data and passed to the subnetworks separately, where the contrastive loss function label Y = 0 if the CXRs both have the ChexPert label “No Finding” label (i.e. no difference between normal lungs) and Y = 1 if one CXR has an abnormal lung label and the other has a “No Finding” label (i.e. difference between lungs). The paired input CXR images were randomly sampled in a manner so that an equal number of Y = 0 and Y = 1 labels were assigned, for both training and validation. We empirically set the number of CXR image pairs sampled per epoch of training and validation at 6400 and 200 image pairs respectively. For both training and validation, each input image is resized to 336 x 336 pixels followed by a center crop to 320 x 320 pixels. This algorithm was implemented using in Python with the PyTorch package, using the Adam optimizer (28) (initial learning rate = 0.00002, β_1_ = 0.9, β_2_ = 0.999). Batch sizes were fixed at 8 for training and validation. Early stopping of training occurred when the validation loss showed no further improvement after 3 training epochs. The model with the lowest validation loss was saved for further training.
**Step 2**. After pre-training on ChexPert data using weak labels, we train the Siamese neural network on the 314 image COVID-19 training set using mean square error (MSE) loss. Each image pair fed to the Siamese neural network results in an output of the Euclidean distance between the final fully connected layers. This Euclidean distance is an abstraction of difference in pulmonary disease severity between the two input CXRs. The “error” of the MSE loss is the difference between the Euclidean distance and the absolute difference in the labeled mRALE scores between the two input images. The input image pairs are randomly sampled during training and validation, with 1600 and 200 image pairs sampled per epoch, respectively. For training, each input image is resized to 336 x 336 pixels followed augmentation with random rotations of ±5° and random crop of 320 x 320 pixels. For validation, each input image is resized to 336 x 336 pixels followed by a center crop to 320 x 320 pixels. This training step was also implemented using the Adam optimizer, with the same hyperparameters as the previous step, and batch sizes of 8. Early stopping was set at 7 epochs without improvement in validation loss. The model with the lowest validation loss was saved for testing evaluation.

#### Occlusion sensitivity maps for visualizing Siamese neural network outputs

To generate an occlusion map, patches of 32 x 32 pixels in the paired input images are occluded (patch area pixel intensities equal the mean of the patch) iteratively across the entire image (stride length 16 pixels). For each iteration, both occluded images are passed through the Siamese neural network and a Euclidean distance is calculated. An increased difference between this Euclidean distance and the non-occluded baseline Euclidean distance indicates that part of the image is important to the network and can be represented as a heat map. When evaluating disease severity in a single image, as in this case, an occlusion sensitivity map is generated for each comparison of the image-of-interest to each image in the pool of ChexPert “No Finding” images. The median of these occlusion sensitivity maps is used for visualization.

#### Statistics

To evaluate differences in patient gender and age in the COVID-19 data sets, we used the Chi-square test and Mann-Whitney test (two-sided), respectively. The associations between variables including the PXS score, mRALE score, and inter-rater mRALE scores were calculated using the Spearman rank correlation (ρ). For evaluating inter-rater reliability of longitudinal CXR change labels, linear Cohen’s kappa (κ) was used. For comparing radiologist agreement with the algorithm on longitudinal change assessment, we used the Spearman rank correlation. For comparison of the PXS score between patients with and without intubation/death, the Mann-Whitney test was used (two-sided). For evaluating the correlation between the time interval between admission and intubation/death, we used the Spearman rank correlation. For area under the receiver operator characteristic curve analysis, bootstrap 95% confidence intervals were calculated. These calculations were performed using the *scipy* and *sklearn* Python packages. The threshold for statistical significance was considered *a priori* to be P<0.05. Data visualizations were performed using the *Seaborn* Python package.

### Supplemental Results

#### Inter-rater correlation in assigning mRALE scores

In the 314-patient COVID-19 training set, the rank correlation between the assigned mRALE score of the two raters was good (ρ=0.84, P<0.001). In the 154-patient COVID-19 internal test set, the rank correlations of the assigned mRALE score between the three raters was similar (ρ=0.82, 0.82, 0.84, P<0.001 in all cases). In the 113-patient COVID-19 external test set, the rank correlations of the assigned mRALE score between the three raters was also similar (ρ=0.85, 0.82, 0.84, P<0.001 in all cases).

## REFERENCES

1. ACR Recommendations for the use of Chest Radiography and Computed Tomography (CT) for Suspected COVID-19 Infection | American College of Radiology. https://www.acr.org/Advocacy-and-Economics/ACR-Position-Statements/Recommendations-for-Chest-Radiography-and-CT-for-Suspected-COVID19-Infection. Accessed March 27, 2020.

2. Rubin GD, Haramati LB, Kanne JP, et al. The Role of Chest Imaging in Patient Management during the COVID-19 Pandemic: A Multinational Consensus Statement from the Fleischner Society. Radiology. Radiological Society of North America; 2020; 201365http://pubs.rsna.org/doi/10.1148/radiol.2020201365. Accessed April 14, 2020.

3. Wong HYF, Lam HYS, Fong AH-T, et al. Frequency and Distribution of Chest Radiographic Findings in COVID-19 Positive Patients. Radiology. Radiological Society of North America; 2019;201160http://pubs.rsna.org/doi/10.1148/radiol.2020201160. Accessed March 27, 2020.

4. M.N. A, L.C. H, M. M, et al. Interobserver reliability of the chest radiograph in community-acquired pneumonia. Chest. 1996;110(2):343-350http://www.ncbi.nlm.nih.gov/pubmed/8697831. Accessed March 29, 2020.

5. Loeb MB, Carusone SBC, Marrie TJ, et al. Interobserver Reliability of Radiologists’ Interpretations of Mobile Chest Radiographs for Nursing Home-Acquired Pneumonia. J Am Med Dir Assoc. Elsevier; 2006;7(7):416-419http://www.ncbi.nlm.nih.gov/pubmed/16979084. Accessed March 29, 2020.

6. Neuman MI, Lee EY, Bixby S, et al. Variability in the interpretation of chest radiographs for the diagnosis of pneumonia in children. J Hosp Med. 2012;7(4):294-298http://www.ncbi.nlm.nih.gov/pubmed/22009855. Accessed March 29, 2020.

7. Li MD, Chang K, Bearce B, et al. Siamese neural networks for continuous disease severity evaluation and change detection in medical imaging. npj Digit Med. Nature Publishing Group; 2020;3(1):48http://www.nature.com/articles/s41746-020-0255-1. Accessed March 29, 2020.

8. Bromley J, Bentz JW, Bottou L, et al. Signature Verification using a “Siamese” Time Delay Neural Network. Int J Pattern Recognit Artif Intell. World Scientific Publishing Company; 1993;07(04):669-688http://www.worldscientific.com/doi/abs/10.1142/S0218001493000339. Accessed June 16, 2019.

9. Hadsell R, Chopra S, LeCun Y. Dimensionality Reduction by Learning an Invariant Mapping. 2006 IEEE Comput Soc Conf Comput Vis Pattern Recognit - Vol 2. IEEE; p. 1735-1742http://ieeexplore.ieee.org/document/1640964/. Accessed June 9, 2019.

10. Irvin J, Rajpurkar P, Ko M, et al. CheXpert: A Large Chest Radiograph Dataset with Uncertainty Labels and Expert Comparison. 2019;http://arxiv.org/abs/1901.07031. Accessed January 4, 2020.

11. Warren MA, Zhao Z, Koyama T, et al. Severity scoring of lung oedema on the chest radiograph is associated with clinical outcomes in ARDS. Thorax. BMJ Publishing Group; 2018;73(9):840–846.

12. Zhou F, Yu T, Du R, et al. Clinical course and risk factors for mortality of adult inpatients with COVID-19 in Wuhan, China: a retrospective cohort study. Lancet. Elsevier BV; 2020;395(10229):1054–1062.

13. Huang G, Liu Z, van der Maaten L, Weinberger KQ. Densely Connected Convolutional Networks. Proc - 30th IEEE Conf Comput Vis Pattern Recognition, CVPR 2017. Institute of Electrical and Electronics Engineers Inc.; 2016;2017-January:2261-2269http://arxiv.org/abs/1608.06993. Accessed March 29, 2020.

14. Zeiler MD, Fergus R. Visualizing and Understanding Convolutional Networks. 2013;http://arxiv.org/abs/1311.2901. Accessed December 7, 2019.

15. Phua J, Weng L, Ling L, et al. Intensive care management of coronavirus disease 2019 (COVID-19): challenges and recommendations. Lancet Respir Med Elsevier; 2020;0(0)http://www.ncbi.nlm.nih.gov/pubmed/32272080. Accessed April 14, 2020.

16. Taylor E, Haven K, Reed P, et al. A chest radiograph scoring system in patients with severe acute respiratory infection: A validation study. BMC Med Imaging. BioMed Central Ltd.; 2015; 15(1).

17. Sheshadri A, Shah DP, Godoy M, et al. ProGression of the Radiologic Severity Index predicts mortality in patients with parainfluenza virus-associated lower respiratory infections. Russell CJ, editor. PLoS One. Public Library of Science; 2018;13(5):e0197418https://dx.plos.org/10.1371/journal.pone.0197418. Accessed April 13, 2020.

18. McClain L, Hall M, Shah SS, et al. Admission chest radiographs predict illness severity for children hospitalized with pneumonia. J Hosp Med. John Wiley and Sons Inc.; 2014;9(9):559–564.

19. Rajpurkar P, Irvin J, Zhu K, et al. CheXNet: Radiologist-Level Pneumonia Detection on Chest X-Rays with Deep Learning. 2017;http://arxiv.org/abs/1711.05225. Accessed January 4, 2020.

20. Lakhani P, Sundaram B. Deep learning at chest radiography: Automated classification of pulmonary tuberculosis by using convolutional neural networks. Radiology. Radiological Society of North America Inc.; 2017;284(2):574–582.

21. Hurt B, Kligerman S, Hsiao A. Deep Learning Localization of Pneumonia. J Thorac Imaging. Ovid Technologies (Wolters Kluwer Health); 2020;1.

22. Liao R, Rubin J, Lam G, et al. Semi-supervised Learning for Quantification of Pulmonary Edema in Chest X-Ray Images. 2019;http://arxiv.org/abs/1902.10785. Accessed January 4, 2020.

23. Kennedy S, Simon B, Alter HJ, Cheung P. Ability of Physicians to Diagnose Congestive Heart Failure Based on Chest X-Ray. J Emerg Med. 2011;40(1):47-52http://www.ncbi.nlm.nih.gov/pubmed/20045607. Accessed January 4, 2020.

24. Hammon M, Dankerl P, Voit-Hohne HL, et al. Improving diagnostic accuracy in assessing pulmonary edema on bedside chest radiographs using a standardized scoring approach. BMC Anesthesiol. BioMed Central; 2014;14:94http://www.ncbi.nlm.nih.gov/pubmed/25364301. Accessed January 4, 2020.

25. Sabottke CF, Spieler BM. The Effect of Image Resolution on Deep Learning in Radiography. Radiol Artif Intell. Radiological Society of North America (RSNA); 2020;2(1):e190015http://pubs.rsna.org/doi/10.1148/ryai.2019190015. Accessed March 29, 2020.

26. Mason D. SU-E-T-33: Pydicom: An Open Source DICOM Library. Med Phys. John Wiley & Sons, Ltd; 2011; 38(6Part10):3493-3493http://doi.wiley.com/10.1118/1.3611983. Accessed April 12, 2020.

27. The OpenCV Library | Dr Dobb’s. https://www.drdobbs.com/open-source/the-opencv-library/184404319. Accessed April 12, 2020.

28. Kingma DP, Ba J. Adam: A Method for Stochastic Optimization. 2014;http://arxiv.org/abs/1412.6980. Accessed June 16, 2019.

